# Three-month treatment with monoclonal antibodies targeting the CGRP pathway is associated with multi-domain improvement of sensory processing and cortical network efficiency: results from a prospective case-control study

**DOI:** 10.1101/2025.05.24.25328294

**Authors:** Lara Klehr, Anne Thiele, Merle Bendig, Christine Kloetzer, Thorsten Herr, Nursena Armagan, Sebastian Strauss, Robert Fleischmann

## Abstract

Monoclonal antibodies targeting calcitonin gene-related peptide (CGRP mAbs) are effective drugs for migraine prevention. Worsening of symptoms following their discontinuation challenges their consideration as disease-modifying migraine drugs (DMMD). This study investigates whether changes in sensory processing and cortical network efficiency under CGRP mAb treatment.

**Methods:** 22 patients suffering episodic migraine (21 female, 46.2±13.8 years) and 22 age-/gender-matched controls received visual and somatosensory evoked potentials (VEPs, SSEPs) assessments, and quantitative electroencephalography (qEEG). Patients were investigated before (V0),after three months (V3), and headache characteristics additionally followed-up at 6 and 12 months, of treatment with CGRP mAbs. Controls were assessed only once.

**Results:** Facilitation of VEP at V0 in patients shifted to habituation at V3 following treatment with CGRP mAbs (Δslope: −0.37±0.83, p=0.03). VEP habituation at V3 did not differ from controls. SSEPs were equally attenuated in patients and controls throughout the study. QEEG parameters in patients indicated impaired network efficiency at V0 that normalized at V3, and were unlike evoked potential studies correlated with six and twelve month outcomes.

**Conclusion/ Interpretation:** Improved cortical network efficiency and sensory processing suggests disease-modifying effects of CGRP mAbs with delayed clinical effects on headache. Relapse after withdrawal may reflect insufficient central adaptation in some patients.

## Introduction

Migraine is a cycling disorder of sensory processing with substantial medical and socio-economic burden to affected individuals and society (1). The availability of monoclonal antibodies (mAbs) targeting the calcitonin gene-related peptide (CGRP) pathway has ushered in a novel era of well-tolerated and effective preventive treatments (2). However, their presumed peripheral mechanism of action i.e. primarily targeting the trigemino-vascular pathway, raises questions about their central disease-modifying capabilities and sustained therapeutic effects following treatment discontinuation (3, 4). Although discontinuation of preventive treatment with CGRP mAbs is recommended in several national guidelines, rapid deterioration of headache frequency and burden in the vast majority of patients after termination was repeatedly shown (5). Few studies investigated markers of central disease activity and pathophysiological mechanisms of migraine in order to elucidate whether or not CGRP mAbs may potentially act as disease-modifying migraine drugs (DMMD), i.e. not only interfere with pain generation but also change fundamental pathomechanisms in the central nervous system. Both hypothalamic activity and dysfunctional brainstem processing of noxious stimuli have been reported to improve during treatment with CGRP mAbs, suggesting potential DMMD properties (6, 7). These studies, however, seem to be in apparent contradiction to clinical results, i.e. the proportion of patients that relapse. Given that migraine is considered a disorder of multimodal sensory processing, lack of sustained efficacy may be resolved by continued deficient higher-order sensory processing despite improved hypothalamic and brainstem function (8). In other words, thalamo-cortical dysfunction may remain unchanged during prevention with CGRP mAbs and underly deterioration. This hypothesis remains to be investigated.

This study thus aims to clarify whether multi-domain sensory processing and cortical network efficiency are affected by preventive treatment with CGRP mAbs. Co-primary endpoints were interictal changes of electrophysiological measures of somatosensory and visual processing in patients affected by episodic migraine, which have frequently been reported as deficient in migraine (9), though not consistently across studies and with considerable interindividual variability (10–12). Further endpoints involve the comparison of sensory processing between individuals with migraine and healthy controls as well as changes of global cortical network efficiency following three-month preventive treatment with CGRP mAbs (13).

## Methods

### Ethical approval and study registration

This study was prospectively registered at clinicaltrials.gov (Identifier: NCT04019496) and approved by the local ethics committee (Identifier: BB 168/18). All procedures adhered to the Helsinki declaration in its latest revision and were conducted in line with current guidelines for good clinical practice (ICH E6(R2)). All patients and controls were provided detailed study information and gave their written consent for the study and the use of their data.

### Study design and participant selection

We enrolled 22 patients suffering episodic migraine and 22 age-/gender-matched controls in this prospective case-control study. Patients with episodic migraine were recruited through the outpatient headache service of a tertiary care university hospital. All patients included in the study had to meet three distinct criteria. The episodic migraine had to be diagnosed using the ICHD-3. Furthermore they needed to fullfil the requirement of failed treatment with four out of five preventive drugs for migraine, such as ß-Blockers, Flunarizine, Amitriptyline, Topiramate and Valproate. Finally, the completion of a three-month migraine diary was mandatory for acceptance into the study. Controls were selected from an internal database maintained by the neurology department of the University Hospital in Greifswald, which comprises individuals such as students, hospital staff, and other volunteers who have previously participated in studies and expressed interest in future research participation. To ensure a gender- and age-matched control group, we additionally recruited students to reflect the age distribution of the younger migraine participants.

Participants were assessed around the same time of the day by the same study personnel. The patients with migraine were tested twice during the study period. It is well established that neurophysiological properties can vary in close temporal proximity to migraine attacks, with changes occurring during the premonitory and postdromal phases. These fluctuations are attack-specific and may not reflect enduring changes in brain function. While it is possible that certain neurophysiological measures remain stable during attacks, the interictal phase is particularly relevant when assessing disease-modifying effects. This phase is assumed to better reflect persistent alterations in sensory processing and brain network activity, which may be influenced by preventive treatment even if attack-related features remain unchanged. Our study therefore focused on capturing neurophysiological properties during the interictal interval, where meaningful changes would indicate an effect of treatment on the underlying disease mechanisms of migraine. Thus, patients were advised not to present for assessments when they suffered a migraine attack within 24 hours before to to ensure that data were collected during the interictal period. Patients suffering chronic migraine were excluded for the same reason. Baseline assessments were conducted before starting the treatment with CGRP mAbs and included the measurement of visual and somatosensory evoked potentials and a 10-minute resting EEG-recording. The first round of tests was marked as V0 and ended with the first injection of CGRP mAbs. Three months later, the patients with migraine repeated the tests which was marked as V3. The healthy controls were investigated only once. Exploratory chart reviews of the headache characteristics were conducted at 6 and 12 months.

### Primary and secondary endpoints

Changes in two domains of sensory processing following three-month preventive treatment with CGRP mAbs in episodic patients with migraine were investigated as co-primary endpoints in this study. We used two different types of stimuli to investigate the multimodal sensory processing.

The visual evoked potentials (VEPs) and the somatosensory evoked potentials (SSEPs). The visual evoked potentials were measured using pattern-reversal stimulation. The patients were seated 1,5m in front of a screen displaying a checkerboard pattern of black and whites squares (alternating with a frequency of 3Hz, an luminance of 50cd/m and a contrast of 80%, the check size was approximately 1 degree of visual angle) in a quiet and dimmed light room. Electrodes were placed in the midline over the occipital region 2,5cm above the inion (Oz) as an active electrode and over the frontal region (Fz) as an reference. Signals were filtered with a 1–100 Hz bandpass. The ground electrode was placed on the arm. The measurements were taken with a frequency of 4kHz. Each eye was tested monocular with 600 stimuli.

Habituation of VEPs was quantified by the standardized slope (beta coefficient) of the linear regression of VEP amplitudes (peak-to-peak from N75 to P100 component) against stimulation order (i.e. values <0 indicating habituation, >0 indicating facilitation of successive stimuli). To calculate this, all trials were averaged offline in MATLAB across six consecutive blocks of 100 stimuli, and the habituation slope was derived from the block-wise amplitudes.

The somatosensory evoked potentials were measured after the stimulation of the median nerve during an EEG recording representing the active electrode and reference electrode. The ground electrode was placed on the arm. Each side was stimulated twice with 500 single stimuli and a one minute break in between. Averaged traces were calculated offline for each run and each side, then averaged across sides to obtain a single SSEP waveform. The frequency of stimulation was 4.4Hz, the frequency of measurement was 2kHz, and signals bandpass-filtered with 30–1000 Hz. The intensity was determined before the test by determining the motor threshold and multiplying it by 1.5. Habituation of SSEPs (N20 component) was calculated in analogy to VEP.

Secondary endpoints included comparisons of VEP and SSEP processing in patients with migraine at baseline and following three-month treatment with CGRP mAbs in comparison to healthy controls.

### Exploratory endpoints

Patients with migraine were shown to be characterised by changes of cortical network efficiency, both in electroencephalographic (EEG) and evoked-potential studies (13, 14). We therefore sought to investigate well-established measures of cortical network physiology using graph theory based on EEG data (15). Resting-state EEG data was acquired using a 64- channel dry EEG channel system (Waveguard touch, ANT Neuro, Enschede, Netherlands). Ten-minutes of EEG data were recorded with eyes closed and subsequently processed in a Matlab environment, which included band-pass filtering (0.5-100Hz), notch filter (50Hz) and manual rejection of artifacts. Remaining 10-second epochs were then split into samples of 4096 data points per channel and imported into BrainWave (version 0.9.151.7.2., University Medical Center Amsterdam, Netherlands (16)). Brainwave yields sensor-level graph characteristics of cortical network efficiency expressed as minimum spanning tree metrics, computed separately for the delta (0.5–3 Hz), theta (4–7 Hz), and alpha (8–13 Hz) frequency bands. Kappa is an indicator of degree distribution and larger values mean that network nodes are more distinctly weighted, which is considered more efficient (17). The Leaf Fraction is the ratio of the number of nodes that have only one edge and a higher leaf fraction indicates better network integration. The Network Diameter gives back the distance between any two nodes ion the network, i.e. larger values are associated with poorer network integration. Tree hierarchy is evaluated as final parameter which is an indicator of the balance between the number of leaf and betweenness centrality, i.e. larger values are considered more beneficial since network efficiency increases while avoiding hub overloads.

### Statistics

Statistical analyses were carried out using the Statistical Package for the Social Sciences (SPSS v25.0, IBM, Armonk, NY, USA). Continuous data were analysed for normal distribution using histogram plots before performing descriptive and inferential statistics. Unless stated differently, normal distribution was confirmed. Descriptive normal distributed data are presented as group means ± standard deviation. When data distribution was non-normal, median values and interquartile ranges (IQR) were presented instead. Inferential comparisons of evoked potential studies between group means were done using paired t-Tests for comparisons between patients at baseline and after three months of treatment. Unpaired t-Tests were used for comparisons between healthy subjects and patients. Inflation of alpha error due to multiple tests assessing the co-primary endpoints was corrected using the Bonferroni method. Other tests were not adjusted given their exploratory character, which needs to be considered in their interpretation. Inferential statistics of cortical network efficiency, i.e. EEG data, were first done using a global test (analysis of variance with network parameters, subject type (i.e. cases, controls) and visits (baseline, three-month follow-up) as factors). Significant results in the global test were post-hoc tested (Tukey’s test) for between group differences.

## Results

We enrolled 22 patients (21 female, mean age 46.2 ± 13.8 years; 4 migraine with aura (MwA)) and 22 matched controls (21 female, mean age 47.6 ± 14.9 years). There were no dropouts in either group. Twelve patients received Erenumab, five patients Galcanezumab and five patients Fremanezumab monthly for an episodic migraine. Headache diaries confirmed that all patients were assessed at least 24 hours after their last migraine attack (distribution illustrated in supplemental figure 1), which in this cohort was a sufficient interval to avoid statistically significant potential carry-over effects from temporally close premonitory or postdromal changes (see supplemental table 1). Diary evaluations furthermore yielded a median headache frequency of 13 (IQR: 10.5 – 14) days/month, mean duration of 8.0 ± 5.3 h/attack and mean headache intensity of 4.9 ± 1.9 at baseline. Following three months of CGRP mAb treatment, headache frequency (5 (IQR: 3-10) days/month, p<0.001 vs. V0) and attack duration (5.4 ± 4.2 h/attack, p=0.014 vs. V0) significantly decreased, while headache intensity remained unchanged (5.1 ± 2.5, p=0.96 vs. V0). Twelve patients achieved a reduction of their headache frequency by ≥50% after 3 months of treatment and were classified responders. Headache frequency further decreased after 6 (5.7 ± 4.1 days/month) and 12 months (4.9 ± 3.3 days/month) of treatment.

### Visual evoked potentials

The slope parameter of the VEP habituation significantly differed between patients with migraine and controls indicating facilitation of subsequent in the migraine group (0.25±0.57 [range: -0.78-1.36]) and habituation in the control group (-0.20±0.89 [range: −1.85-1.34], p= 0.031). VEP habituation significantly changed from facilitation into habituation in the migraine group after three months of treatment (-0.12±0.85 [range: −1.52-2.08], p=0.030), which was not significantly different from controls (p= 0.40). Results are summarized in figure 1.

**Figure 1.**
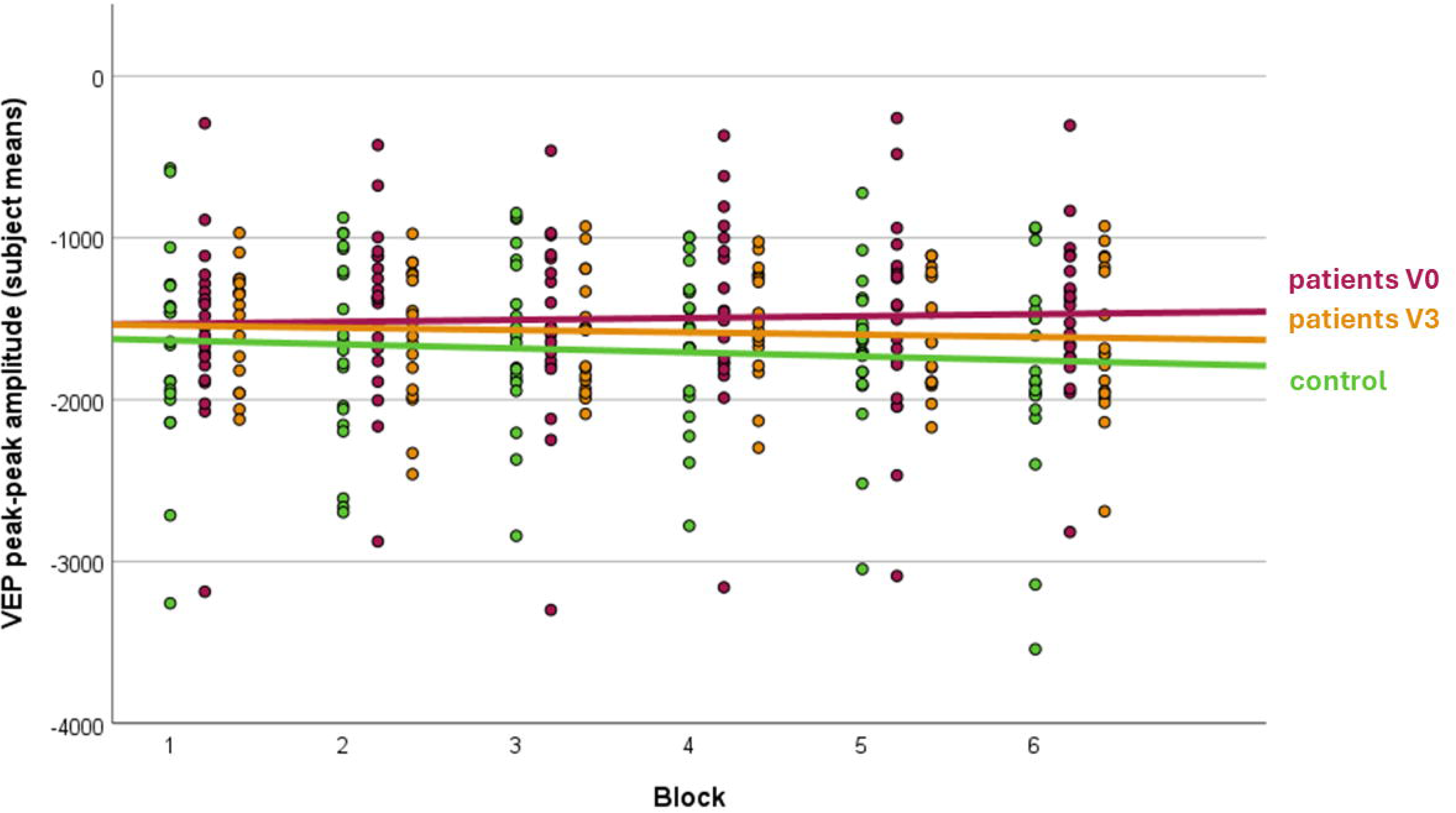
VEP amplitudes in controls and migraine patients before and during CGRP monoclonal antibody treatment. The figure displays the average peak-to-peak amplitudes of visual evoked potentials (VEPs) per stimulus block across three conditions: healthy controls, migraine patients before treatment, and migraine patients three months after initiation of CGRP mAb therapy. Fit lines are shown for each group to illustrate the amplitude trends across blocks. While overall amplitude levels are comparable between groups, the migraine group at baseline exhibits a clear pattern of facilitation over successive blocks, in contrast to the habituation observed in healthy controls and in patients following treatment. These trends indicate that habituation deficits at baseline are normalized after three months of preventive CGRP-targeted therapy.

### Somatosensory evoked potentials

SSEP amplitudes remained stable across the treatment interval in migraine patients and did not differ significantly from controls at either baseline or follow-up. In patients, the mean peak-to-peak amplitude was 641.01±1239.31µV at V0 and 651.91±1245.61µV three months after initiation of CGRP monoclonal antibody therapy (V3). This numerical increase was not statistically significant (p=0.36). In between-subject comparisons, SSEP amplitudes in patients did not differ from healthy controls at baseline (p=0.97) or after treatment (639.41±134.51µV; p=0.822).There was also no significant difference of SSEP habituation between patients with migraine and control subjects at baseline, with findings indicating habituation in both groups (migraine: −1.13±0.16 [range: −1.48--0.86], controls: −1.12±0.15 [range: −1.44--0.86], p=0.43; figure 2). The habituation of SSEPs in patients with migraine assessed at three-month follow-up (-1.09±0.12 [range: −1.34--0.91]) was unchanged as compared to baseline (p=0.19) and remained indifferent from control subjects (p=0.41).

**Figure 2.**
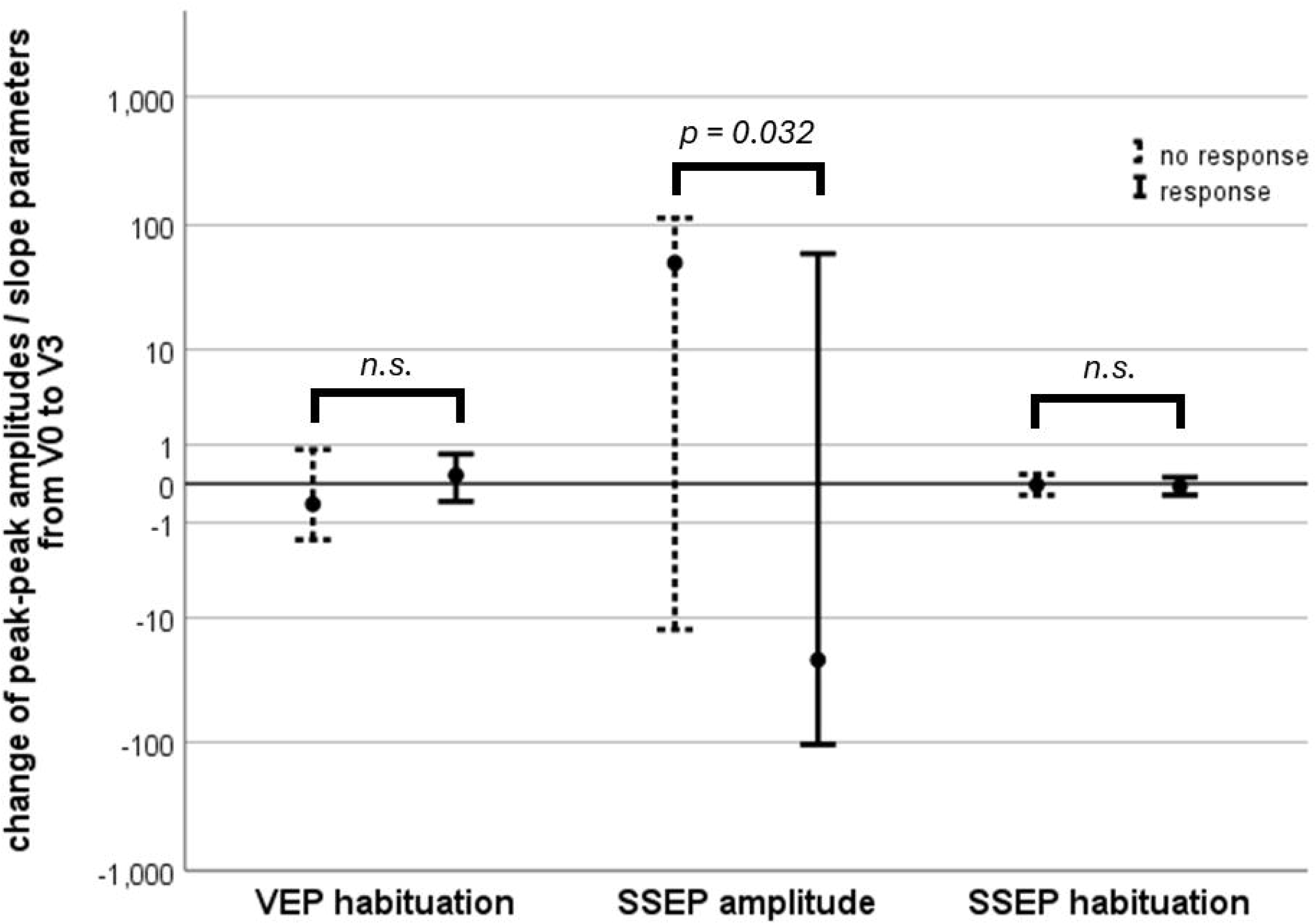
Association between changes in neurophysiological markers and clinical treatment response. The figure illustrates the relationship between changes in habituation slopes of visual (VEP) and somatosensory (SSEP) evoked potentials, as well as changes in SSEP amplitudes, and the clinical response to treatment in migraine patients. Mean changes from baseline (V0) to follow-up (V3) are shown. While habituation changes did not significantly differ between responders and non-responders for either modality, a significantly greater reduction in SSEP amplitudes was observed in patients who responded to treatment after three months of CGRP monoclonal antibody therapy.

**Figure 3.**
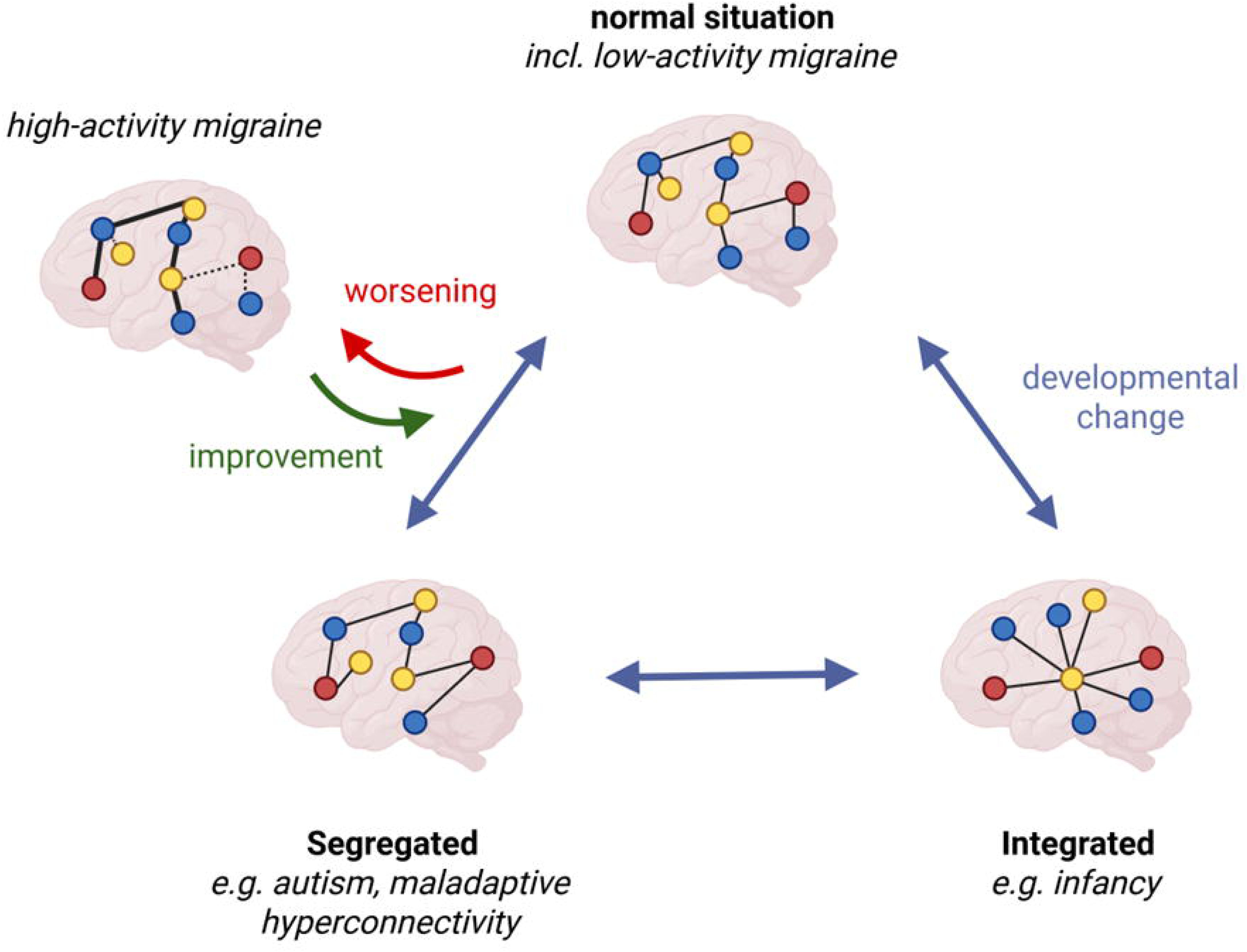
Schematic representation of sensor-level connectivity changes in migraine with and without preventive treatment. Simplified brain network organizations are depicted across developmental stages, including how findings in migraine patients align with graph- theoretical network models. In patients who did not receive preventive treatment, the network diameter was larger, and kappa values were lower, indicating a more segregated and less resilient network architecture. After three months of treatment with CGRP mAbs, network parameters became indistinguishable from those of matched control subjects.

### Relationship between changes of neurophysiological parameters and treatment response

We conducted correlation analyses between changes in headache frequency after 3 months of treatment and alterations in neurophysiological measures, including evoked potentials and resting-state connectivity. Neither changes in SSEP or VEP nor any connectivity measures were significantly correlated with changes in monthly headache days. We then performed binary comparisons between treatment responders and non-responders after 3 months, defining responders as those with a ≥50% reduction in monthly headache days. Again, network connectivity measures showed no significant association with treatment response. However, evoked potential studies revealed a significantly greater decrease in SSEP amplitudes among responders while changes of habituation did not differ between groups (figure 2).

Exploratory correlation analyses were performed to assess changes in headache frequency at 6 and 12 months of treatment with CGRP mAbs. Changes in evoked potential measures at three months showed no significant correlation with long-term changes in headache frequency (all p>0.05). However, connectivity analyses revealed a significant association between a decrease in headache frequency and an increase in the leaf fraction in the theta band (r=0.42, p =0.042), suggesting that a less segregated network is linked to improved long-term migraine activity. Additionally, there were trends indicating that an increase in kappa in the theta band (r=-0.31, p=0.093) and a decrease in diameter in the delta band (r=0.32, p=0.092) may also be associated with a reduction in headache frequency. Both findings further support the notion that a less segregated network is related to improved migraine outcomes.

### Resting-state connectivity

An overview of the descriptive data of all network parameters across frequency bands can be found in table 1, table 2 and table 3. Minimum spanning tree parameters assessed at V0 significantly differed between the migraine group and controls regarding Kappa (F=5.6, p=0.02), which was not frequency-specific (migraine: 6.6±2.7, controls: 7.2±2.6). There was furthermore a significant frequency-specific difference regarding the Diameter at baseline (F=6.5, p=0.002). Post-hoc testing revealed that the Diameter in the migraine group was larger in the alpha band (migraine: 0.56±0.14, controls: 0.46±0.10, p=0.03). Kappa (F=2.89, p=0.15) and Diameter (F=1.60, p=0.21) assessed in patients with migraine were not significantly different from control values after three months of treatment (schematic of connectivity changes in figure 3). Neither tree hierarchy nor leaf fraction significantly differed between patients and controls at any time or frequency band (all p>0.05).

## Discussion

This study identified significant improvements of sensory processing and brain network efficiency in patients with migraine following three months of treatment with CGRP mAbs, which normalized in comparison with control subjects. These mechanisms are linked to migraine pathophysiology apart from structures directly associated with the generation of pain in migraine attacks. Restitution of VEP habituation and impaired brain network efficiency indicates improved multi-modal information processing, which is understood to be key to interictal mechanisms in migraine pathophysiological leading to recurrent migraine attacks. Evidence for their amelioration in this study supports the notion of disease-modifying properties of the treatment with CRPS mAbs.

### Changes of evoked-potential studies in relation to previous findings

Migraine is considered a disorder of multi-domain sensory processing, which involves attenuation of evoked-potential amplitudes but also lack of habituation of subsequent stimuli (8, 18). We were able to replicate previous findings of impaired sensory habituation in patients with migraine in our study, which is well in line with existing literature. However, it is important to acknowledge that other well-controlled studies, including blinded protocols, have not replicated this effect (10, 12). Furthermore, substantial interindividual variability has been reported, even among healthy controls, calling into question the robustness of habituation as a reliable diagnostic marker on the individual level (11). Our own control group also showed a wide range of VEP slopes, including facilitation, reinforcing the importance of interpreting group-level effects with caution. Nonetheless, Ambrosini et al. also showed the significant lack of habituation in interictal episodic patients with migraine in comparison to healthy controls (19). Furthermore, they proved that the combination of altered auditory and visual evoked potentials significantly increases the likelihood of migraines in diagnostic process (20). A different study put focus on somatosensory evoked potentials. Restuccia et al. measured the habituation of high frequency oscillations and N20 SSEP components in patients with migraine and healthy controls. They demonstrated the impaired habituation in N20 response in patients with migraine which underlies changes depending on the migraine state (21). Other studies focussed on intervention effects on visually and somatosensory evoked potentials. For example, di Lorenza et al. discovered a restored habituation of VEPs and SSEPs in patients with migraine after a ketogenic diet which depended on clinical improvement (22). Coppola et al. showed the effects of repetitive transcranial magnetic simulation on somatosensory evoked potentials (23). They also stated the lack of somatosensory habituation in patients with migraine compared to healthy controls and restored habituation in patients with migraine after 10Hz rTMS. Moreover, they were able to show an increase of high-frequency somatosensory oscillations between attacks which reflects the thalamo-cortical activity. Extending on these previous studies, our study is the first to demonstrate that deficient VEP habituation can be restored through treatment with CGRP mAbs that do not have the ability to directly interfere with visual information processing in the central nervous system (3). SSEP habituation was found to be unimpaired in patients with migraine throughout the study, which is a finding that differs from previous studies despite using an established protocol (24). Differences in patient cohorts, migraine cycle, clinical fluctuations and generally larger heterogeneity of SSEP habituation than other sensory domains potentially underly this discrepancy (25).

### Changes of cortical network efficiency in relation to previous findings

In this study we not only found changes of cortical sensory processing but also improvements of brain network efficiency in patients with migraine. Impaired brain network efficiency before treatment with CGRP mAbs was restored to levels comparable with controls subjects, which is a novel finding. While previous studies have evaluated the impact of CGRP mAbs on brain network connectivity and sensory processing, they used different study designs that did not include control groups, and employed distinct network connectivity measures. Specifically, La Rocca et al. and de Tommaso et al. both examined Galcanezumab treatment and its effects on brain connectivity and visual evoked potentials (26, 27). Both studies reported that Galcanezumab significantly altered cortical processing, particularly within visual networks, leading to improvements in network integration and sensory reactivity. Our findings similarly demonstrate an overall increase in network efficiency following CGRP mAb treatment, with restoration of VEP habituation and a reduction in network diameter in the alpha band, suggesting improved cortical communication. Notably, while our study examined global sensor-level connectivity at rest, La Rocca et al. reported regional changes in the occipital and frontal areas during visual stimulation.

There are only a few previous studies that used EEG in patients with migraine. De Tommaso et al. found that light-induced EEG changes differed interictally by migraine subtype. Patients without aura showed alpha synchronization and reduced connectivity, while those with aura showed beta desynchronization and increased connectivity (28). Formisano et al. investigated the effect of flunarizine on cortical physiology and showed that EEG abnormalities normalized along with migraine frequency in patients with migraine (29).

In line with the argumentation related to changes of evoked potential findings, it is not expected that CGRP mAbs directly interfere with information processing on a cortical level. Yet, absence of repeated migraine attacks may indirectly ameliorate the interaction of thalamico-cortical loops and enhance metabolic support of neurons through more efficient sensory processing, both of which enable more efficient network function (8, 13).

### Neurophysiological findings and disease modification

The discontinuation of preventive treatment with CGRP mAbs often leads to worsening of migraine activity, which contradicts the notion of disease modification through CGRP mAbs.

Our findings, however, clearly show that there are far-reaching improvements of migraine pathophysiology even after three months of treatment, which are remote from the site of action of CGRP mAbs and supports DMMD properties. This being said, not all patients experience a rebound to their previous migraine activity but improvement persists over months, especially in those patients that best responded regarding their migraine frequency (30). It is thus possible that identified improvements in sensory processing and brain network efficiency are driven by patients that will remain with a low migraine activity even after discontinuation of the preventive medication while others with lesser improvement of neurophysiological parameters will experience a rebound. This interpretation is strongly supported by the finding that exploratory clinical data at 6 and 12 months showed partial correlations with improvements in network efficiency after 3 months of treatment. This suggests that network changes might contribute to disease-modifying effects, which are reflected in long-term migraine activity (4). In contrast, more direct measures of sensory processing, such as evoked potential studies, appear to reflect immediate improvements related to migraine attacks. Specifically, VEPs are linked to the processing of visual information (e.g., phonophobia), while SSEPs are associated with pain processing. This divergence between sensory modalities is not unprecedented. A recent study reported that treatment with a CGRP mAb normalized abnormal SSEP habituation in migraine patients (24). Their cohort showed deficient SSEP habituation at baseline that improved with treatment, whereas our cohort displayed normal SSEP habituation prior to treatment but a reduction in SSEP amplitudes in responders following three months of treatment in our cohort. These differences may reflect sample heterogeneity, disease phase, or baseline neurophysiological state. Together, these findings support the idea that CGRP-targeted therapy can induce central neurophysiological changes in migraine, though the degree and detectability of such effects may depend on modality, timing, and patient characteristics. Thus, our findings do not contradict the clinical course in a proportion of patients but may explain why some patients have a lasting benefit while others deteriorate, i.e. the duration of treatment with CGRP mAbs until disease modification is achieved may vary between patients. This hypothesis needs to tested in prospective studies that also explore changes after discontinuation of the treatment.

## Limitations

Our findings offer insights into a potential aspect of migraine pathophysiology and disease activity but we did not assess clinical changes of interictal symptoms. This being said, improvement of interictal migraine can only be inferred based on neurophysiological data but its direct association to the migraine phenotype requires confirmation in future studies, e.g. reduction of interictal migraine burden. We also cannot rule out that we identified spontaneous fluctuations within a three-month cycle and behavioural adaptions to the experimental setup since healthy control subjects were only assessed once. It is possible that their responses would have changed over time. While our own data show that facilitation patterns can also occur in healthy individuals, the group-level effect we observed supports the interpretation that the changes in VEP responses are specific to the treatment of migraine. Although facilitation of VEPs has frequently been reported in migraine, it is not consistently observed in all patients or across all studies. Finally, the attentional demands and duration of VEP recordings may have introduced variability in responses and contributed to a more pronounced habituation in controls, who may have been less focused during later blocks.

The lack of a placebo group or follow-up examinations of the control group does limit the interpretation of effects specific to CGRP mAbs adjusted for natural fluctuations and non-pharmacological treatment effects. This should be acknowledged in future trials.

## Conclusions

In this study we identified the normalization of cortical network efficiency and sensory processing after a three month treatment with CGRP mAbs. Alterations in sensory processing and cortical network efficiency were detectable after 3 months of CGRP mAb treatment, but correlations with headache frequency emerged only at 6 and 12 months. This suggests that central changes occur earlier, while their translation into clinical effects on headache frequency requires sustained treatment, likely reflecting indirect effects of peripheral CGRP blockade. Deterioration after withdrawal may be explained by the lack of identified disease-modifying effects on higher-order sensory processing in some patients.

## Data Availability

All data produced in the present study are available upon reasonable request to the authors

## Acknowledgment

Not applicable.

## Conflicts of Interest and Disclosure

The Author(s) declare(s) that there is no conflict of interest

## Authors contributions

Conception and design of the study: RF, LK. Review of methods: RF, SS, NA. Electrophysiological data acquisition and pre-processing: AT, LK, MB, NA, TH. Electrophysiological data evaluations: RF, NA, TH, SS. Clinical data acquisition: RF, SS, CK. Interpretation of results: RF, LK. Drafting the manuscript: LK, RF. All authors reviewed and approved the final version of manuscript.

## Funding

This research was supported by the Soyka-Förderpreis für Schmerzforschung der Novartis Pharma GmbH Deutschland. CS was supported by the DFG Clinician Scientist Program.

## Article Highlights

- Migraine is often associated with deficient habituation of visual evoked potentials, though findings vary between individuals and across studies
- Brain network efficiency is impaired in migraine as compared to controls
- Three-month treatment with CGRP mAbs restores VEP habituation and brain network function rendering them comparable to controls
- Findings indicate that treatment with CGRP mAbs contributes to disease modification outside the pain-related structures

## Notes

### Competing Interest Statement

The authors have declared no competing interest.

### Clinical Protocols

https://clinicaltrials.gov/study/NCT04019496

### Funding Statement

This research was supported by the Soyka-Foerderpreis fuer Schmerzforschung der Novartis Pharma GmbH Deutschland. CS was supported by the DFG Clinician Scientist Program.

### Author Declarations

Ethikkommission an der Universitaetsmedizin Greifswald Institut fuer Pharmakologie Felix-Hausdorff-Str. 3 17487 Greifswald Identifier: BB 168/18

