## supplemental figure 1 for "Three-month treatment with monoclonal antibodies targeting the CGRP pathway is associated with multi-domain improvement of sensory processing and cortical network efficiency: results from a prospective case-control study"

**Supplemental figure 1,** occurrence of migraine attacks before and after measurement at baseline. There were no migraine attacks within 24 hours of assessments.

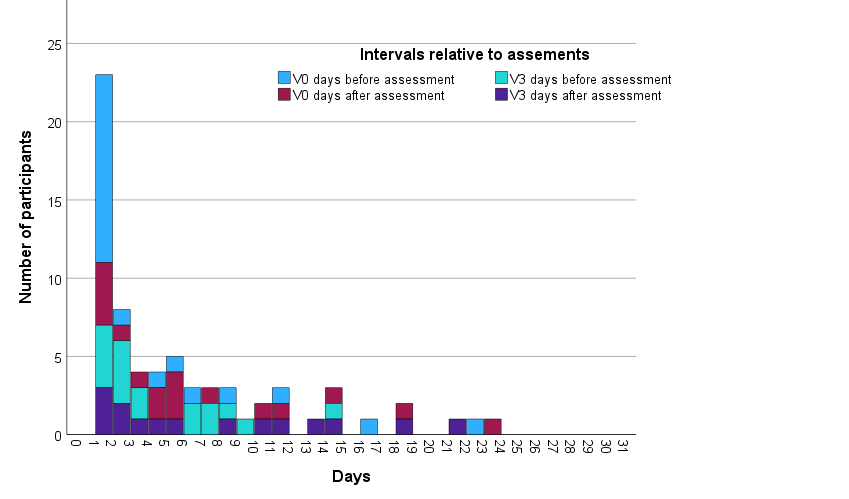

**Supplemental table 1,** multivariate test of the statistical effect of assessment times to or from the next migraine attack on sensory processing.

| Effect | Value | F | Hypothesis df | Error df | Sig. |
| --- | --- | --- | --- | --- | --- |
| Intercept | .995 | 130.746^b^ | 6.000 | 4.000 | <.001 |
| ATKPREV0 | .532 | .758^b^ | 6.000 | 4.000 | .638 |
| ATKPOSV0 | .482 | .620^b^ | 6.000 | 4.000 | .714 |
| ATKPREV3 | .427 | .497^b^ | 6.000 | 4.000 | .788 |
| ATKPOSV3 | .722 | 1.734^b^ | 6.000 | 4.000 | .309 |

Design: Intercept + ATKPREV0 + ATKPOSV0 + ATKPREV3 + ATKPOSV3

ATKPREV0 = days migraine attack occurred before assessment at V0

ATKPOSV0 = days migraine attack occurred after assessment at V0

ATKPREV3 = days migraine attack occurred before assessment at V3

ATKPOSV3 = days migraine attack occurred after assessment at V3
