## Supplementary material for "Three-month treatment with monoclonal antibodies targeting the CGRP pathway is associated with multi-domain improvement of sensory processing and cortical network efficiency: results from a prospective case-control study": table 1

**Table 1. MST parameters in the delta band.**

|  | | **MST Kappa** | **MST Diameter** | **MST Leaf Fraction** | **MST Tree hierarchy** |
| --- | --- | --- | --- | --- | --- |
| **Baseline** | | | | | |
| migraine | Mean | 3.0221 | .2225 | .5311 | .3946 |
|  | Std. Deviation | .34729 | .04694 | .04215 | .05318 |
| control | Mean | 4.0794 | .2018 | .5983 | .4342 |
|  | Std. Deviation | 1.51373 | .07323 | .07306 | .06388 |
| **3-month follow-up** | | | | | |
| migraine | Mean | 5.9637 | .1841 | .7079 | .4218 |
|  | Std. Deviation | 3.41416 | .08403 | .18222 | .08640 |
