## Supplementary material for "Three-month treatment with monoclonal antibodies targeting the CGRP pathway is associated with multi-domain improvement of sensory processing and cortical network efficiency: results from a prospective case-control study": table 2

**Table 2. MST parameters in the theta band.**

|  | | MST Kappa | MST Diameter | MST Leaf Fraction | MST Tree hierarchy |
| --- | --- | --- | --- | --- | --- |
| **Baseline** | | | | | |
| migraine | Mean | 8.3196 | .4008 | .4722 | .3634 |
|  | Std. Deviation | .93896 | .11430 | .10758 | .09007 |
| control | Mean | 8.4580 | .5000 | .4950 | .3679 |
|  | Std. Deviation | 1.30315 | .08165 | .07619 | .04777 |
| **3-month follow-up** | | | | | |
| migraine | Mean | 9.8294 | .3774 | .6448 | .4197 |
|  | Std. Deviation | 3.32601 | .16142 | .22045 | .07960 |
