## Supplementary material for "Three-month treatment with monoclonal antibodies targeting the CGRP pathway is associated with multi-domain improvement of sensory processing and cortical network efficiency: results from a prospective case-control study": table 3

**Table 3. MST parameters in the alpha band.**

|  | | MST Kappa | MST Diameter | MST Leaf Fraction | MST Tree hierarchy |
| --- | --- | --- | --- | --- | --- |
| **Baseline** | | | | | |
| migraine | Mean | 8.5092 | .5588 | .5051 | .3381 |
|  | Std. Deviation | 1.07409 | .13974 | .08995 | .09467 |
| control | Mean | 9.0091 | .4600 | .4850 | .3340 |
|  | Std. Deviation | 1.54446 | .10488 | .07835 | .06127 |
| **3-month follow-up** | | | | | |
| migraine | Mean | 8.5089 | .4522 | .5596 | .3702 |
|  | Std. Deviation | 3.19379 | .17578 | .22516 | .12956 |
